# Prevalence of Post COVID-19 Condition among healthcare workers: self-reported online survey in four African countries

**DOI:** 10.1101/2023.06.22.23291768

**Authors:** Hager Elnadi, Ahmad I. Al-Mustapha, Ismail A. Odetokun, AbdulAzeez Adeyemi Anjorin, Rasha Mosbah, Folorunso O. Fasina, Youssef Razouqi, Kwame Sherrif Awiagah, Jean Baptiste Nyandwi, Zuhal E. Mhgoob, George Gachara, Mohamed Farah Yusuf Mohamud, Bamu F. Damaris, Aala MohamedOsman Maisara, Mona Radwan

**Author notes:** Corresponding author: Ahmad I. Al-Mustapha (,).

## Abstract

The impact of Post COVID-19 Condition is ongoing despite the WHO declaration that the pandemic has ended. In this study, we explore the prevalence of PCC among healthcare workers (HCWs) in four African Countries and its influence on their professional performance. This study was conducted as an online cross-sectional survey of healthcare workers from four African countries (Cameroon, Egypt, Nigeria, and Somalia) between the 20^th^ of December 2021 to 12^th^ of January 2022. We determined the prevalence of PCC based on the WHO case definition and assessed variables associated with a higher prevalence of PCC in these countries using univariable and multivariable logistic regression analyses. A total of 706 HCWs from four African countries were included in this survey. Most of the HCWs were aged between 18-34 years (75.8%, n=535). Our findings showed that 19.5% (n=138) of the HCWs had tested positive for SARS-CoV-2. However, 8.4% (n=59) were symptomatic for COVID-19 but tested negative or were never tested. Two-thirds of the HCWs (66.4%, n=469) have received a COVID-19 vaccine and 80.6% (n=378) of those vaccinated had been fully vaccinated. The self-reported awareness rate of PCC among the HCWs was 16.1% (n=114/706) whereas the awareness rate of PCC among COVID-19-positive HCWs was 55.3 % (n=109/197). The prevalence of PCC among HCWs was 58.8% (n=116). These changes include the self-reported symptoms of PCC which included headache (58.4%, n=115), fatigue (58.8%, n=116), and muscle pain (39.6%, n=78). Similarly, 30% (n=59) and 20.8% (n=41) of the HCWs reported the loss of smell and loss of taste long after their COVID-19 infection, respectively. Some HCWs (42%, n=83) believed that their work performance has been affected by their ongoing symptoms of PCC. There was no significant difference in the prevalence of PCC among the vaccinated and unvaccinated HCWs (p > 0.05). Of the socio-demographic variables, age (older HCWs between 45-54 years; OR:1.7; 95% CI: 1.06, 10.59; p = 0.001) and location (Egypt; OR:14.57; 95% CI: 2.62, 26.76; p = 0.001) were more likely to have experienced PCC than other age groups and countries respectively. The study revealed low prevalence of PCC among the surveyed HCC. In addition, it observed the need for adequate medical and psychological support to HCWs with PCC, improve their COVID-19 vaccination uptake, and conduct mass advocacy campaigns on PCC.

**Graphical abstract:** COVID-19 positivity rate (n =197), vaccination rate (n =706), PCC awareness rate (n = 114), and prevalence of PCC (n = 116) in HCWs across four African countries.

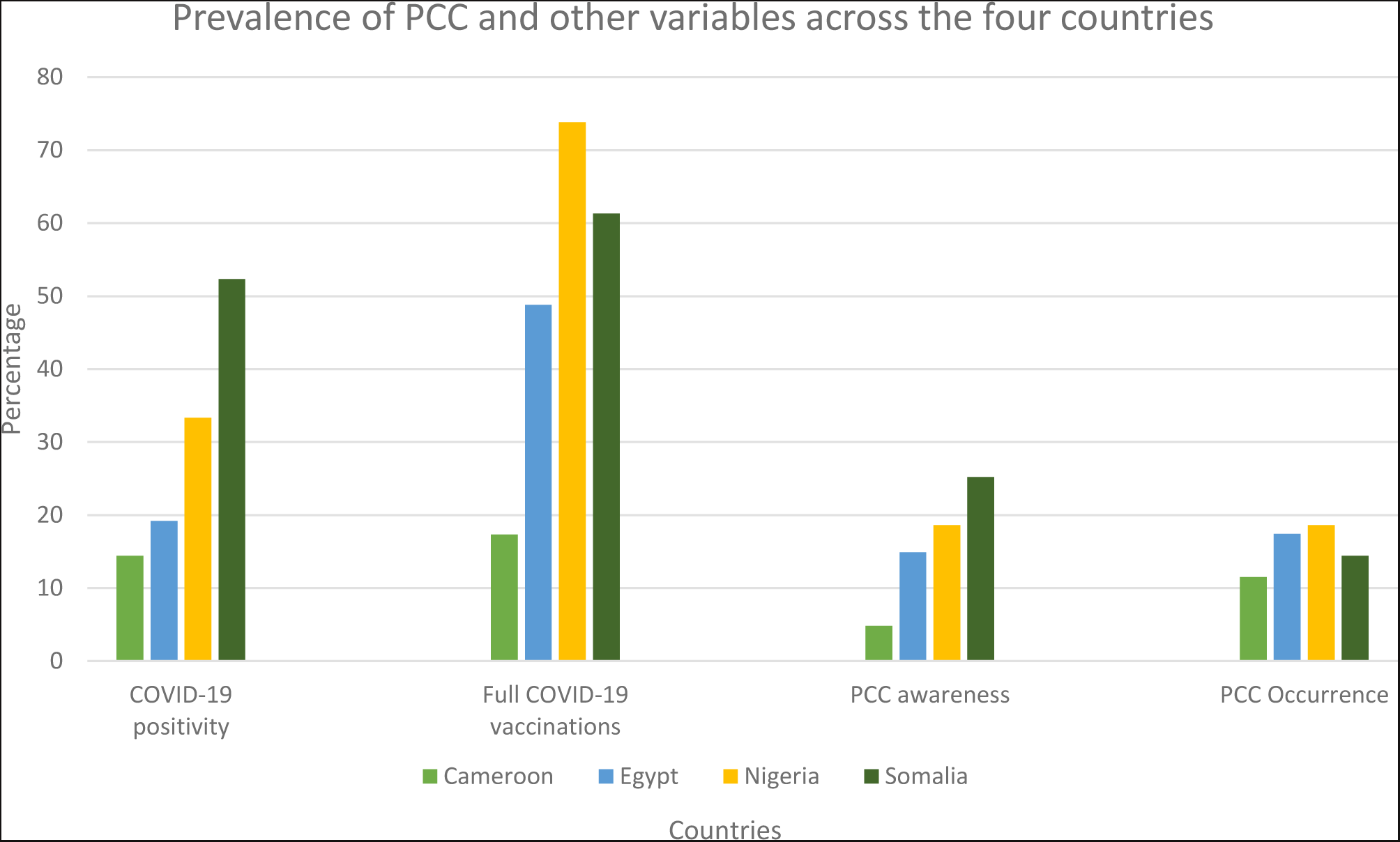

## 1. Introduction

The severe acute respiratory syndrome coronavirus 2 (SARS-CoV-2) is the causative agent of the 2019 coronavirus disease (COVID-19) pandemic. SARS-CoV-2 mainly affects the respiratory system and the disease, COVID-19 has various clinical presentations, which vary from mild to severe, especially among elderly and immunocompromised patients [1].

By 7^th^ of June 2023, more than 767 million confirmed COVID-19 cases have been reported, which has resulted in 6,941,095 million deaths globally. In Africa, COVID-19 reported cases were around 9,534 million with 175,373 COVID-19-associated deaths (case fatality rate of 2%) [2]. To curb the spread of the SARS-CoV-2 virus, governments across the world initially instituted non-pharmaceutical interventions (the use of face masks, hand hygiene, physical distancing, and related measures) and later pharmaceutical interventions (mainly vaccines). The accelerating and efficient development, production, distribution, and acceptance of the COVID-19 vaccines have helped curb the disease spread, reduce hospitalization rates, and reduced the severity of the disease, particularly among infected patients [3]. The tremendous effect of the COVID-19 vaccines in reducing the disease transmission and the return of routine daily life worldwide, led to WHO’s announcement on 5^th^ of May 2023, that COVID-19 pandemic is no longer a public health emergency [4].

After a COVID-19 infection, most of the ill patients return to their normal health status within days to a few weeks. However, some patients continue to suffer from ongoing, recurrent, or long-term health issues post-infection [5,6]. For instance, some patients were reported to experience a wide range of symptoms such as headache, fatigue, change in taste or smell, irregular menstruation, and mood changes among others, for months, post-infection [7–8].

To gain a significant understanding of these long-term COVID-19 adverse events, the World Health Organization (WHO) published a temporary clinical case definition for such cases, [5,9]. In October 2021, WHO announced that symptoms/signs which were previously termed long-term COVID-19, chronic COVID-19, or Post COVID-19 syndrome, are now identified as Post COVID-19 Condition (PCC) [5]. Despite the identification of these COVID-19 sequelae as PCC by WHO, they lack clear classifications and several things remain unclear [5]. For instance, the impact of vaccinations on the incidence of PCC requires further studies. Al-Aly et al.,[10] reported that COVID-19 vaccines reduced the likelihood of PCC in people who had been infected by only about 15% whereas Antonelli et al., reported that two doses of the COVID-19 vaccines halved the risk of PCC [11].

For instance, PCC had been reported in COVID-19 survivors regardless of their COVID-19 severity (mild or severe), presentation (acute, sub-acute, or chronic), and whether the patient was hospitalized or not [8,9]. In addition, the PCC is similar to the post-SARS (severe acute respiratory syndrome coronavirus 1) and the Middle East Respiratory Syndrome Coronavirus (MERS CoV) syndromes (with symptoms such as fatigue, myalgia, psychiatric presentation, as well as pulmonary and bone complications) that have affected SARS, MERS CoV survivors for up to 4-15 years post-infection [12–18].

Given the insufficient information about its effects on individuals’ health during the illness, or long after recovery, PCC may have a negative impact not only on the patient’s health but also on their personal and professional life. Thus, the USA considered PCC a disability under the Americans with Disabilities Act (ADA) since July 2021 [4]. However, there is a paucity of reliable data about the COVID-19 pandemic and PCC in Africa. Therefore, this study assessed the prevalence of PCC and its association with the work performance of healthcare workers (HCWs) in four African countries.

## 2. Materials and Methods

### 2.1 Ethical clearance

The ethical clearance for this study was obtained from the Kwara State Ministry of Health, Ilorin, Nigeria with reference number MOH/KS/EHC/777/502 as well as the ethical review board of the Faculty of Human Medicine of the University of Zagazig, Egypt (Reference number: ZU-IRB #9241/2-1-2022). We obtained written informed consent from each respondent after brief information on the purpose of the study was provided to them. To participate in the study, a respondent must tick the consent box in the mobile application (ODK). Participation in this survey was voluntary and without prejudice, as participants could withdraw from the survey at any time.

### 2.2 Study Participants and survey methodology

This study presented the preliminary findings from a larger study that was conducted as a cross-sectional online survey of HCWs across Africa between the 20^th^ of December 2021 to 12^th^ of January 2022. HCWs included anyone that provides health services and advice based on formal training and experience. Hence, they included physicians, nurses, veterinarians, etc. The questionnaire was designed using Google Forms (Google incorporated) and administered via online social media platforms such as WhatsApp, Facebook, and E-mails. Our inclusion criteria were age (18 years and above), location (Africa), and occupation (healthcare worker). Other African countries (n=12) with less than 100 respondents were excluded from this preliminary analysis.

### 2.3 Study variables

This study evaluated four self-reported variables among HCWs in each country. These were: 1). COVID-19 positivity rate, 2). COVID-19 full vaccination rate, 3). awareness of PCC, and 4). prevalence of PCC among HCWs. The COVID-19 positivity rate among HCWs was essential to understand the prevalence of COVID-19 in HCWs and evaluating the occupational exposure of HCWs in these countries. The second study variable evaluated the vaccination coverage of HCWs in these countries. Although HCWs were designated as priority recipients for the COVID-19 vaccination, acceptance of the vaccine is voluntary. Hence, our interest in the vaccination status of HCWs. The third variable evaluated the awareness of PCC (especially its symptoms and clinical presentation). This is crucial to the diagnosis of PCC among patients. The final study variable evaluated the prevalence of PCC among HCWs. Based on the WHO case definition, a respondent was classified as having PCC if the individual had a history of probable or confirmed SARS-CoV-2 infection, usually 3 months from the onset of COVID-19, with symptoms lasting for at least 2 months that could not be explained by an alternative diagnosis [5]. So, only respondents that fit the four criteria of the WHO case definition of PCC were regarded as having PCC.

### 2.4 Questionnaire design

The awareness of PCC among HCWs was assessed using a semi-structured pre-validated questionnaire. The survey instrument was further validated by three independent academic examiners to ascertain the content and face validity of the adapted questionnaire as well as observe for any technical glitches. In addition, the reliability of the survey instrument was assessed using the Cronbach Alpha test (with a score of 0.72) based on 11 purposefully selected questions. Finally, the questionnaire was pre-tested among 10 HCWs from each of the four countries before the deployment of the final version for data collection. The results of the pre-test were not included in the final analysis.

The questionnaire was designed in three of the most common languages in Africa (Arabic, English, and French). The back-to-back translation was validated by two linguists to ensure that the intended meaning of each question was not lost. In each of the translations, the questionnaire was divided into 4 sections: a). Socio-demographic information on HCWs b). history and course of COVID-19 infection c). Awareness of PCC among HCWs d). Impact on work performance.

### 2.5 Data analysis

The data obtained from this survey were analyzed using Statistical Package for Social Sciences (SPSS) version 26 (IBM Corp., Armonk, N.Y., USA). We conducted descriptive statistics and summarized the information as frequency and percentages. Chi-square analysis was used to test for association between the four key study variables (COVID-19 positivity rate, COVID-19 full vaccination rate, awareness of PCC, and the prevalence of PCC) in the four countries. Finally, the significant variables (p-value < 0.05) were entered into a logistic regression model (univariable and multivariable) to determine the association between the socio-demographic variables (age, gender, occupation, and their country of origin) and the outcome variable (prevalence of PCC in HCWs). The odds ratios generated from the multivariable logistic regression analysis were used for all the inferences in this study.

## 3. Results

### 3.1 Healthcare worker demographics

A total of 713 HCWs filled out the survey instrument out of which 99% (n=706) gave consent and responded to the questionnaire (Table 1). The distribution from the four African countries was Egypt (n = 281), Nigeria (n = 210), Somalia (n = 111), and Cameroon (n = 104). Most of the HCWs were aged between 18-34 years (75.8%, n=535). Of the respondents, nurses represented 36.3% (n=256) and more female respondents were recruited (55.8%, n=394).

**Table 1.** Demographics of HCWs recruited into this study (n=706).

| Variables | Number (%) |
| --- | --- |
| Nationality |  |
| Cameroon | 104 (14.7) |
| Egypt | 281 (39.8) |
| Nigeria | 210 (29.8) |
| Somalia | 111 (15.7) |
| Occupation |  |
| Dentist | 38 (5.4) |
| Nurse | 256 (36.3) |
| Physician | 143 (20.3) |
| Pharmacist | 42 (6.1) |
| Laboratory technician | 74 (6.7) |
| Veterinarian | 79 (11.2) |
| Others* | 74 (6.7) |
| Age |  |
| 18 - 24 | 283 (40.1) |
| 25-34 | 252 (35.7) |
| 35-44 | 125 (17.7) |
| 45-54 | 36 (5.1) |
| >55 | 10 (1.4) |
| Gender |  |
| Female | 394 (55.8) |
| Male | 301 (42.6) |
| Prefer not to say | 11 (1.6) |
| Do you have a known chronic disease? |  |
| No | 646 (91.5) |
| Yes | 60 (8.5) |

*Others\*: scientific officers, health information technologists, and clinical medical students who work or study in a health facility.*

### 3.2 COVID-19 infection among healthcare workers

Of the 706 HCWs included in this survey, 19.5% (n=138/706) tested positive for SARS-CoV-2 by real-time polymerase chain reaction (RT-PCR), rapid diagnostic test (RDT), or reverse transcription loop-mediated isothermal amplification (RT-LAMP). In addition, 8.4% (n=59/706) of the HCWs were probable COVID-19 cases. During their COVID-19 infection, 22.3% of them had moderate to severe symptoms and were admitted to a health facility (Table 2). Our findings showed that two-thirds of the HCWs (66.4%, n=469) in these countries have received a COVID-19 vaccine and 80.6% (n=378) of those vaccinated had received their second dose (fully vaccinated). A quarter of the HCWs (27.5%, n=38) were tested less than a month before this survey.

**Table 2.** History of COVID-19 infection among healthcare workers recruited into this study.

| Variables | Number (%) |
| --- | --- |
| 1. Have you ever been diagnosed with COVID-19? (n = 706) |  |
| No | 568 (80.5) |
| Yes | 138 (19.5) |
| 2. Have you noticed the symptoms of COVID-19 which were not confirmed (probable cases)? (n = 568) |  |
| No | 509 (72) |
| Yes | 59 (8.4) |
| 3. When was your last COVID-19 test conducted? (n = 127) |  |
| < 1 month ago | 38 (27.5) |
| 2-3 months | 13 (9.4) |
| 3-6 Months | 20 (14.5) |
| 6-12 months | 30 (21.7) |
| 12-18 months | 24 (17.4) |
| >18 months | 2 (2.2) |
| 4. Were you admitted to a health facility during your COVID-19 infection? (n = 197) |  |
| No | 153 (77.7) |
| Yes | 44 (22.3) |
| 5. Have you received any of the approved COVID-19 vaccines? (n = 706) |  |
| No | 237 (33.6) |
| Yes | 469 (66.4) |
| 6. Are you fully vaccinated against COVID-19? (n = 469) |  |
| No | 91 (19.4) |
| Yes | 378 (80.6) |

The awareness rate of PCC among the general HCWs was 16.1% (n=114/706) whereas the awareness rate of PCC among COVID-19-positive HCWs was 55.3 % (n=109/197). Based on the WHO case definition, more than half of the COVID-19-positive HCWs (58.8%, n=116/197) were deemed to have PCC and one-third (35.5%, n=70/197) of them noticed changes in their health status due to the PCC. These changes include the symptoms of PCC which included headache (58.4%, n=115), fatigue (58.8%, n=116), and muscle pain (39.6%, n=78). Similarly, 30% (n=59) of the HCWs reported the loss of smell after their COVID-19 infection and 20.8% (n=41) of the HCWs experienced the loss of taste long after their COVID-19 infection. Other less frequent COVID-19 symptoms experienced by HCWs in the 4 African countries included rash, pins and needles, and memory problems (Figure 1). Half of the HCWs (53.3%, n=105) reported that their symptoms of PCC were mostly mild, and only 6.9% of them (n=14) were admitted due to their PCC or its symptoms (Table 3).

**Figure 1.**
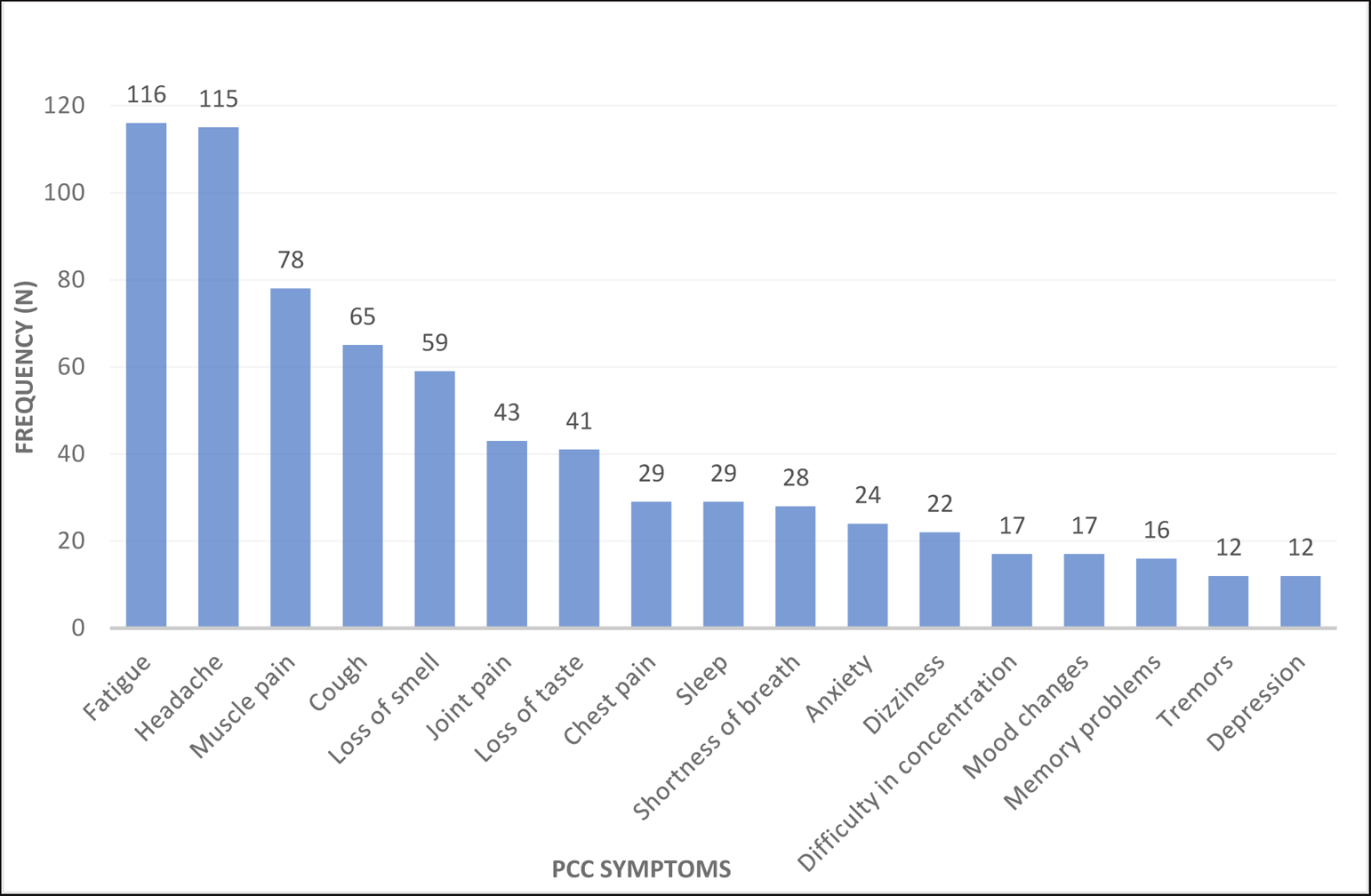
Frequency of PCC symptoms among COVID-19-positive healthcare workers from four African countries, 2022 (n = 197/706).

**Table 3.**
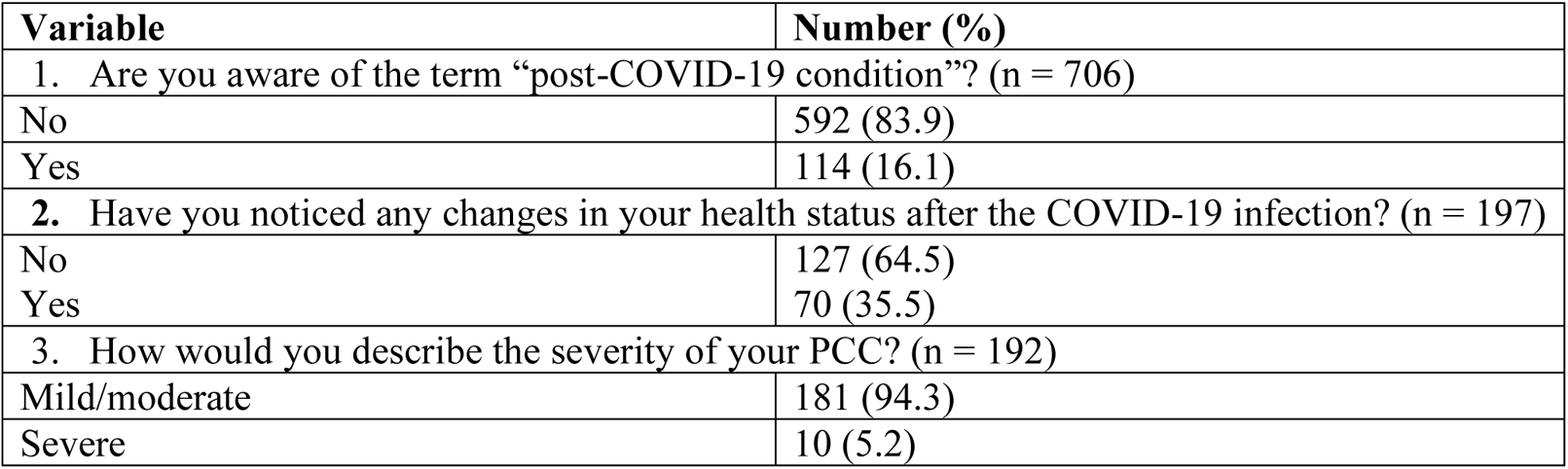
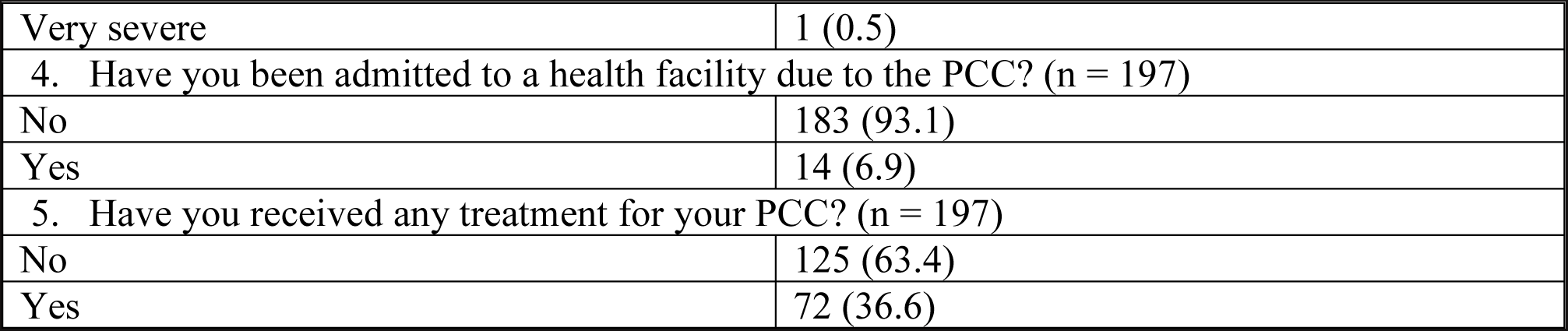
Structure of PCC among HCWs in four African countries (n=197).

| Variable | Number (%) |
| --- | --- |
| 1. Are you aware of the term “post-COVID-19 condition”? (n = 706) |  |
| No | 592 (83.9) |
| Yes | 114 (16.1) |
| 2. Have you noticed any changes in your health status after the COVID-19 infection? (n = 197) |  |
| No | 127 (64.5) |
| Yes | 70 (35.5) |
| 3. How would you describe the severity of your PCC? (n = 192) |  |
| Mild/moderate | 181 (94.3) |
| Severe | 10 (5.2) |
| Very severe | 1 (0.5) |
| 4. Have you been admitted to a health facility due to the PCC? (n = 197) |  |
| No | 183 (93.1) |
| Yes | 14 (6.9) |
| 5. Have you received any treatment for your PCC? (n = 197) |  |
| No | 125 (63.4) |
| Yes | 72 (36.6) |

### 3.3 Work Performance

Only 29.7% (n=58) of the HCWs who experienced COVID-19 and PCC described their work environment as more stressful. In addition, 42% (n=83) of them believed that their performance at work has been affected by their PCC. Approximately half of the HCWs (49.3%, n=97) got tired faster than usual whereas 16.2% of them (n=32) got more forgetful after their COVID-19 infection (Table 4).

**Table 4.**
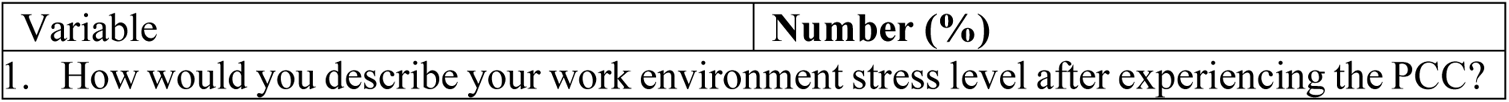
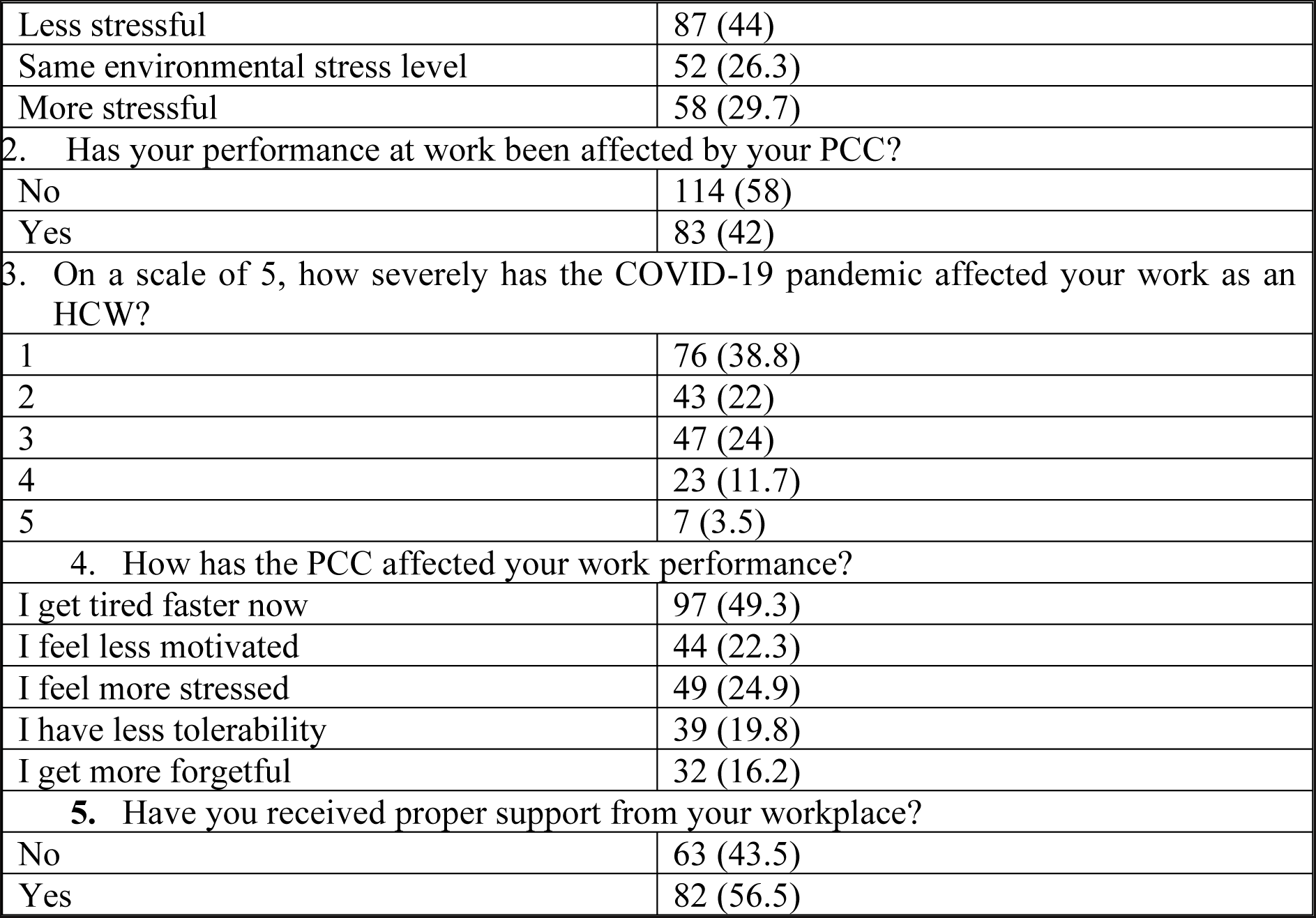
Perception of COVID-19 infection and PCC on work performance among HCWs (n=197).

| Variable | Number (%) |
| --- | --- |
| 1. How would you describe your work environment stress level after experiencing the PCC? |  |
| Less stressful | 87 (44) |
| Same environmental stress level | 52 (26.3) |
| More stressful | 58 (29.7) |
| 2. Has your performance at work been affected by your PCC? |  |
| No | 114 (58) |
| Yes | 83 (42) |
| 3. On a scale of 5, how severely has the COVID-19 pandemic affected your work as an HCW? |  |
| 1 | 76 (38.8) |
| 2 | 43 (22) |
| 3 | 47 (24) |
| 4 | 23 (11.7) |
| 5 | 7 (3.5) |
| 4. How has the PCC affected your work performance? |  |
| I get tired faster now | 97 (49.3) |
| I feel less motivated | 44 (22.3) |
| I feel more stressed | 49 (24.9) |
| I have less tolerability | 39 (19.8) |
| I get more forgetful | 32 (16.2) |
| 5. Have you received proper support from your workplace? |  |
| No | 63 (43.5) |
| Yes | 82 (56.5) |

### 3.4 Post-Covid Condition among African healthcare workers

Across the four countries, there were statistically significant differences in the positivity rate of COVID-19 infection among HCWs, their vaccination rates, their awareness of PCC, and the prevalence of PCC among HCWs (Table S2). HCWs in Somalia tested more positive for COVID-19 than HCWs in other countries (p < 0.05). However, more HCWs in Nigeria (73.8%, n=155) had received full COVID-19 vaccines than in the three other countries (p < 0.05). The highest awareness rate of PCC was in Somalia HCWs where 25.2% of them knew about PCC (Table S2). The prevalence of PCC among COVID-19-positive HCWs was 58.8% (n=116). However, there were no significant differences in the prevalence of PCC among the vaccinated and un-vaccinated HCWs (p > 0.05).

Of the sociodemographic variables, only nationality significantly impacted the prevalence of PCC among HCWs in Africa. Hence, HCWs in Egypt were more likely (OR:14.57; 95% CI: 2.62, 60.76; p = 0.001) to have experienced PCC than HCWs in the three other countries (Table 5). This is further evident in the fact that 90.7% (n=49) of the 54 Egyptian HCWs who tested positive for the SARS-COV-2, experienced PCC.

**Table 5.**
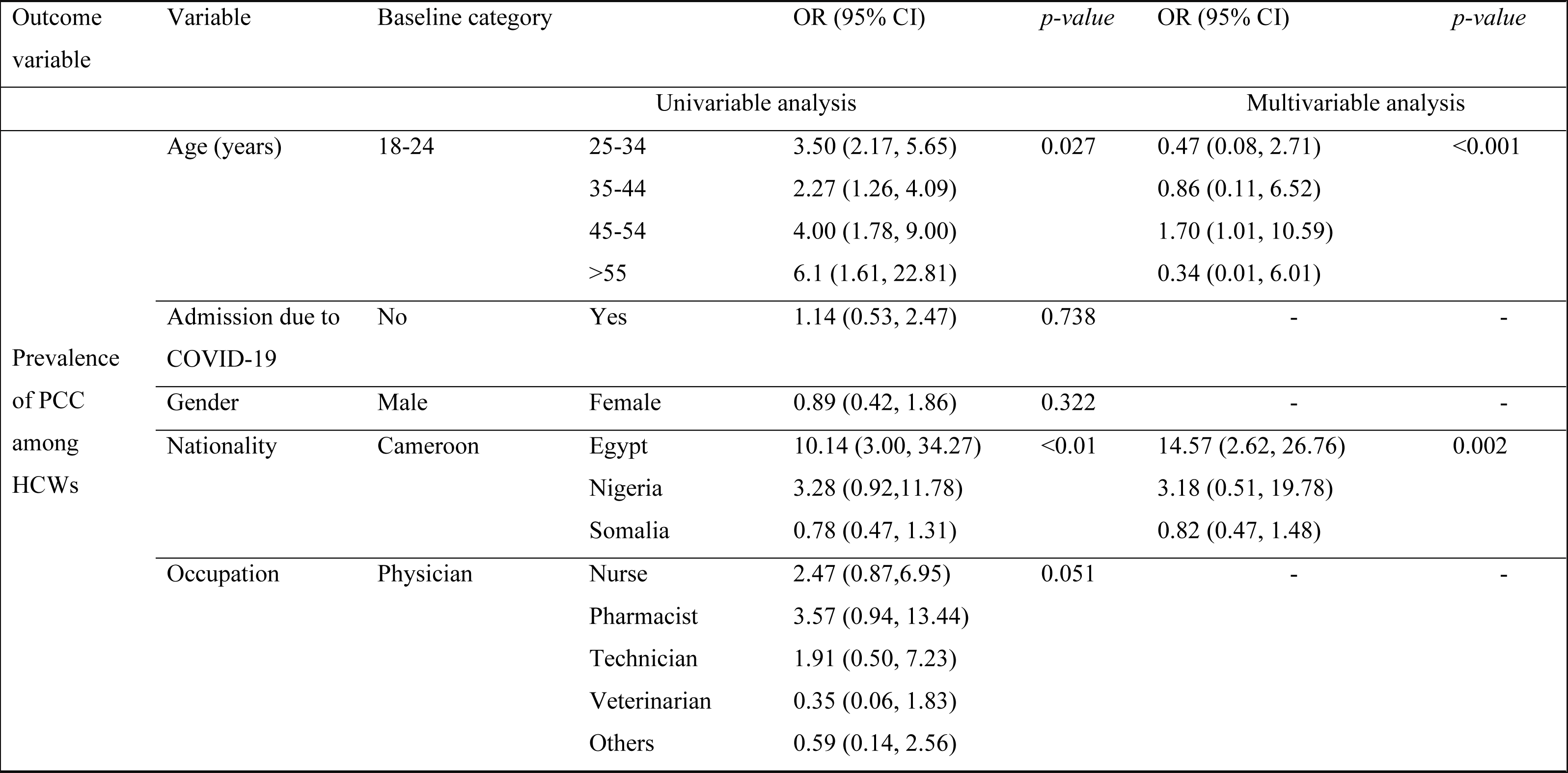
Logistic regression analysis of demographic variables that affected the prevalence of PCC among HCWs.

## 4. Discussion

Globally, HCWs play vital roles as frontline response personnel during the COVID-19 pandemic, especially in emergency responses, public education, and case management. This however exposes them to a higher risk of COVID-19. One of the problems associated with the COVID-19 infection is the persistence of symptoms which is generally referred to as “long COVID” or “PCC” [5]. Here, we present the first PCC survey that targeted HCWs across four African countries (Cameroon, Egypt, Nigeria, and Somalia). The COVID-19 pandemic affected the healthcare system in most African countries.

Our findings showed that 19.5% of HCWs who participated in this study had confirmed the COVID-19 diagnostic test, while 8.4% had COVID-19 symptoms and signs but without a confirmatory test. Therefore, the prevalence of SARS-CoV-2 infection among the HCWs was 27.9% (n=197) in the aforementioned four African countries. The highest COVID-19 positivity rate was among Somali HCWs (52.1%).

Our results showed a higher prevalence rate than recorded by studies such as those of Galanis et al., [17] and Gómez-Ochoa et al., [18], where they observed the seroprevalence of SARS-CoV-2 in African HCWs were around 8.2% and 7-11% in 2021 respectively. However, another study in the same year observed a wider range of prevalence (0 - 45.1%) of COVID-19 among HCWs in 11 African countries [19]. A recent meta-analysis reported that 10.1% of COVID-19 patients were healthcare workers (HCWs) [20]. The variability in the prevalence rate of COVID-19 among HCWs could be attributed to the sensitivities and specificities of the diagnostic tests used (self-reported vs serosurveillance), the study design, the wave of the pandemic in each country, the period the samples were taken, and the country-specific contextual differences. For instance, Muller et al [19] solely relied on rapid antibody diagnostic tests and their results differed between African countries, with 45.1% of seroprevalence in Nigeria and 1.3% seropositive prevalence in Egypt by the end of 2021.

Our findings showed that among the 197 HCWs who suffered from COVID-19, 58.8% of them experienced PCC based on the clinical symptoms and the WHO clinical case definition. This PCC prevalence rate is in line with the reports of Osikomaiya et al. [21] who reported a prevalence of 40.9% for PCC among the general populace in Lagos, Nigeria in 2020. In the same vein, a systematic review of 57 studies with more than 250 000 participants by Groff et al. [22], reported a PCC prevalence of 54% among recovered COVID-19 patients in 2021. Also, Taquet et al. et al. [23] reported that the prevalence of PCC in the USA was 57% during the first 6 months after a positive COVID-19 test in the year 2021.

In addition, Hyassat D, et al., [24] reported that 59.3% of the healthcare providers in Jordan reported more than one persisting COVID-19 symptom, and among them 97.5%, 62.6% and 40.9% reported more than one COVID-19 symptom at 1–3, 3–6 and 6–12 months, respectively, after the acute phase of the infection. This finding is in consonant with our findings. However, Menges et al. [25] reported a much lower PCC prevalence of 26% within 6 to 8 months in COVID-19-positive patients. The slightly higher prevalence of PCC in older health professionals (45-54 years old) than in other age groups could be attributed to the fact that older persons were usually more symptomatic than others [26].

Like other studies across the globe, the most reported symptoms of PCC in this study were fatigue, headache, malaise, headache, myalgia, cough, and loss of smell among other symptoms. These symptoms have been the most common among patients diagnosed with PCC across the globe. For instance, Montenegro et al. [27] and Iwu et al. [28] reported that fatigue, anosmia, headache, and dyspnoea have been the most complaints among PCC patients. In addition, Osikomaiya et al. [23] observed that 12.8% of patients with PCC in Nigeria had persistent fatigability and another 12.8% had a headache after their discharge from the hospital. In the same vein, fatigue was the most reported symptom among Jordanian HCWs [24]. These results opined that most PCC manifestations were ongoing COVID-19 symptoms.

Our findings showed that a large proportion of the HCWs who participated in this study had no prior knowledge about PCC as only 16.6% of them were aware of PCC. For instance, Somali HCWs had the highest awareness rate (25.2%) than the other three countries whilst HCWs from Cameroon had the least awareness rate of PCC as only 4.8% (n=5/104) of them were aware of PCC or its symptoms. The differences in the awareness rate of PCC could be due to the differences in the health systems of each country, the HCW training schedule of each country, and the occupation of the HCWs. However, the PCC awareness rate was high among COVID-19-positive HCWs as 55.3% of them were aware of PCC.

The COVID-19 vaccination rate was different in the four countries. Study participants from Cameroon had the lowest vaccination rate as only 17.3% of them were vaccinated. These low vaccination rates could be because Cameroon has the lowest national COVID-19 vaccination rates of the four countries (Table S1). Although our findings showed that HCWs from Nigeria had the highest vaccination rate, Egypt has the highest national COVID-19 vaccination rate with over 71.8 million vaccinations and 29.3% of its populace fully vaccinated. These results were supported by a previous study in 34 African countries which showed that only 63% of their study participants were willing to accept the COVID-19 vaccine [31].

More longitudinal research are needed to evaluate the impact of vaccinations on the incidence of PCC. Our data showed that the vaccination status had no impact on the prevalence of PCC as there was no significant difference (p > 0.05) in the fully vaccinated, partially vaccinated, and un-vaccinated HCWs. This is contrary to the findings of several studies which reported that COVID-19 vaccines reduced the likelihood of PCC in COVID-19 patients [32]. In the same vein, the category of the HCW (dentist, nurses, physicians, laboratory technicians, veterinarians, and others) was not associated with the prevalence of PCC in the four countries.

Besides the clinical manifestations of COVID-19, several studies have reported the psychological burden of the current pandemic on HCWs irrespective of their SARS-CoV-2 infection status [33–38]. While HCWs know that their profession poses a high infection risk, most of them were afraid to transmit the disease to their families [39]. Furthermore, other studies have observed anxiety, stress, exhaustion, and depression, with increasing rates of burnout among HCWs in Africa especially due to the insufficiency of personal protective equipment and the limited number of standardized care facilities in most African countries [39–41].

Similar to the findings of these studies, our findings showed that psychological symptoms were experienced by HCWs in the four African countries included in this study. For instance, 12.2%, 8.6%, and 6.1% of the HCWs had anxiety issues, mood changes, and depression respectively. This psychological burden could affect the perceived stress level in HCWs. Previously, Salazar et al. [42], Schwartz et. al. [43], and Chew et. al. [44] reported that the stress level at health facilities during the pandemic was considered more apparent than pre-pandemic era, with rising levels of burnout syndrome. Conversely, the majority of the HCWs felt the work environment have been either less stressful (44%) or no change in the work stress level (26.3%) during the pandemic compared to the pre-pandemic era, and only 29.7% of the HCWs felt higher stress level after the pandemic at their workplaces.

This general perception among the general HCWs was different from the PCC-affected HCWs, as 42% of the latter felt that because of their PCC, their work performance has been negatively affected. For instance, 49.3% of them got easily fatigued, while others were more stressed (24.9%), and lost their enthusiasm (22.3%). In an international study involving 56 countries, Davis et al. [45] reported that 45.2% of their study participants (n=3762) required a reduced workload due to ongoing symptoms of PCC. In addition, Twycross, A. [46] opined that the current support scheme for healthcare professionals with PCC in the UK requires immediate review.

Despite the several challenges of each country’s healthcare system, HCWs in this study opined that their respective work institutions provided their affected professionals with appropriate work support when they got back to work as reported by 56.5% of our HCWs that had PCC. Recently, several studies have discussed the importance of modifying the health services settings in Africa to deliver better care for COVID-19 patients and especially for healthcare workers in African countries [39, 47–53]. Interestingly, several African counties are making remarkable strides to handle PCC in Africa [21]. These strides started with the recognition of PCC, its burdens, as well as its management and control. In addition, Egypt recently established the first clinic dedicated to PCC patients’ care and treatment [54].

This study has several limitations. Firstly, the sample size is small and the findings should not be generalized for each country. In addition, online, self-reported surveys could be biased and usually skewed toward the younger, urban population with access to the internet. In addition, since the prevalence of PCC was based on self-reports, there was a high likelihood of misclassification, misunderstanding of some questions, and over-estimation of the prevalence of PCC, especially in the probable COVID-19 cases. Despite the limitations of our survey methodology (which makes it impossible to generalize for the HCWs in each country), we believe that this study will provide baseline information on PCC among HCWs in Africa.

## 5. Conclusion

This study presented the COVID-19 positivity rate, vaccination rate, awareness of PCC, and prevalence of PCC among HCWs in four African countries. Generally, HCWs were among the frontline response team during the COVID-19 pandemic. Therefore, it is essential to protect HCWs with a focus on improved mental health and psychological well-being and the development of national programmes for the occupational health and safety of HCWs [55]. The COVID-19 positivity rate was high in Nigeria and Somalia. Hence, infection, prevention, and control measures as well as regular training should be instituted for HCWs in these countries. The very low COVID vaccination rate in Cameroon is worrying and the relevant health authorities in Cameroon must improve vaccination uptake, especially among HCWs. Similarly, the awareness of PCC was lowest among HCWs from Cameroon. So, mass advocacy campaigns on PCC and its varying implications should be provided for the general public in Cameroon with an emphasis on HCWs. Finally, HCWs from Egypt had the highest prevalence of PCC. Hence, adequate medical and psychological support should be provided.

## Data Availability

All data produced in the present study are available upon reasonable request to the authors

## Declarations

- Ethics approval and consent to participate The ethical approval for this study was obtained from the ethical review board (ERB) of the Kwara State Ministry of Health, Ilorin, Nigeria (reference number MOH/KS/EHC/777/502) as well as the ERB of the Faculty of Human Medicine of the University of Zagazig, Egypt (Reference number: ZU-IRB #9241/2-1-2022). This study was conducted in accordance with the World Medical Association’s Declaration of Helsinki and written informed consent was obtained from each respondent.
- Consent for publication Not Applicable
- Availability of data The datasets used and/or analyzed during the current study are available from the corresponding author upon reasonable request.
- Competing interests The authors declare that they have no competing interests.
- Funding No funding was received for this study.
- Authors’ contributions HE, AIA, IAO, AAA, RM, BFD, MR, and MFYM collected data from these countries. HE and AIA did the statistical analysis and wrote the draft manuscript. All authors read and approved the final version of the manuscript.

## Acknowledgements

We acknowledge Dr Abubakar Musa Imam for reviewing the manuscript.

## Legend

**Table S1.** COVID-19 vaccination status in the four African countries as of 6^th^ June.

| Country | Population | Total doses given | Fully vaccinated |
| --- | --- | --- | --- |
| Cameroon | 26.6 million | 3.66 million | 3.11 million |
| Egypt | 102 million | 56.91 million | 42.34 million |
| Nigeria | 206 million | 112.5 million | 69.79 |
| Somalia | 15.9 million | 8.37 million | 7.04 million |

**Table S2.** Association between the nationality of HCWs and their COVID-19 positivity, vaccination rate, and awareness of PCC.

| Variables | Egypt<br>(n=281) | Nigeria<br>(n=210) | Cameroon<br>(n=104) | Somalia<br>(n=111) | $\chi^2$ | p-value |
| --- | --- | --- | --- | --- | --- | --- |
| COVID-19 positivity (n=197) | 54 (19.2) | 70 (33.3) | 15 (14.4) | 58 (52.3) | 13.1 | 0.001 |
| Full COVID-19 vaccination (n=378) | 137 (48.8) | 155 (73.8) | 18 (17.3) | 68 (61.3) | 9.3 | 0.025 |
| Awareness of Post-COVID-19 condition (n=114) | 42 (14.9) | 39 (18.6) | 5 (4.8) | 28 (25.2) | 5.3 | 0.002 |
| Prevalence of PCC (n=116) | 49 (17.4) | 39 (18.6) | 12 (11.5) | 16 (14.4) | 21.2 | 0.001 |

$\chi$ – Chi-square

## Notes

### Competing Interest Statement

The authors have declared no competing interest.

### Funding Statement

This study did not receive any funding

### Author Declarations

The ethical clearance for this study was obtained from the Kwara State Ministry of Health, Ilorin, Nigeria with reference number MOH/KS/EHC/777/502 as well as the ethical review board of the Faculty of Human Medicine of the University of Zagazig, Egypt (Reference 134 number: ZU-IRB #9241/2-1-2022). We obtained written informed consent from each respondent after brief information on the purpose of the study was provided to them. To participate in the study, a respondent must tick the consent box in the mobile application (ODK). Participation in this survey was voluntary and without prejudice, as participants could withdraw from the survey at any time.

## References

1. Desai AD, Lavelle M, Boursiquot BC, Wan EY. Long-term complications of COVID-19. Am J Physiol Cell Physiol. 2022 Jan 1;322(1):C1–C11. doi: 10.1152/ajpcell.00375.2021.

2. WHO Coronavirus (COVID-19) Dashboard. WHO Coronavirus (COVID-19) Dashboard with Vaccination Data. Available online: https://covid19.who.int/ (accessed on 8 March 2022).

3. Eroglu B, Nuwarda RF, Ramzan I, Kayser V. A Narrative Review of COVID-19 Vaccines. Vaccines (Basel). 2021;10(1):62. Published 2021 Dec 31. doi:10.3390/vaccines10010062

4. WHO (2023). *Coronavirus disease (covid-19) pandemic*, *World Health Organization*. Available at: https://www.who.int/europe/emergencies/situations/covid-19 (Accessed: 15 June 2023).

5. CDC. National Center for Immunization and Respiratory Diseases (NCIRD), Division of Viral Diseases, https://www.cdc.gov/coronavirus/2019-ncov/long-term-effects/index.html. Accessed on 22 February 2022

6. WHO. A Clinical Case Definition of Post COVID-19 Condition by a Delphi Consensus, 6 October 2021. Available online: https://www.who.int/publications/i/item/WHO-2019-nCoV-Post_COVID-19_condition-Clinical_case_definition-2021.1 (accessed on 8 December 2021).

7. Pavli A, Theodoridou M, Maltezou HC. Post-COVID Syndrome: Incidence, Clinical Spectrum, and Challenges for Primary Healthcare Professionals. Arch Med Res. 2021;52(6):575–581. doi:10.1016/j.arcmed.2021.03.010

8. Yong SJ. Long COVID or post-COVID-19 syndrome: putative pathophysiology, risk factors, and treatments. Infect Dis (Lond). 2021 Oct;53(10):737–754. doi: 10.1080/23744235.2021.1924397.

9. Lechner-Scott J, Levy M, Hawkes C, Yeh A, Giovannoni G. Long COVID or post COVID-19 syndrome. Mult Scler Relat Disord. 2021 Oct;55:103268. doi: 10.1016/j.msard.2021.103268.

10. Al-Aly Z, Bowe B, Xie Y. Long COVID after breakthrough SARS-CoV-2 infection. Nature Medicine. 2022;28(7):1461–1467.

11. Antonelli M, Penfold RS, Merino J, Sudre CH, Molteni E, Berry S, Canas LS, Graham MS, Klaser K, Modat M, Murray B. Risk factors and disease profile of post-vaccination SARS-CoV-2 infection in UK users of the COVID Symptom Study app: a prospective, community-based, nested, case-control study. The Lancet Infectious Diseases. 2022 Jan 1;22(1):43–55.

12. Zhao FC, Guo KJ, Li ZR.. Osteonecrosis of the femoral head in SARS patients: seven years later. Eur J Orthop Surg Traumatol. 2013;23(6):671–677. [PubMed] [Google Scholar]

13. Das KM, Lee EY, Singh R, et al. Follow-up chest radiographic findings in patients with MERS-CoV after recovery. Indian J Radiol Imaging. 2017;27(3):342–349. [PMC free article] [PubMed] [Google Scholar]

14. Lee SH, Shin H-S, Park HY, et al. Depression as a Mediator of Chronic Fatigue and Post-Traumatic Stress Symptoms in Middle East Respiratory Syndrome Survivors. Psychiatry Investig. 2019;16(1):59–64. [PMC free article] [PubMed] [Google Scholar]

15. Rogers JP, Chesney E, Oliver D, Pollak TA, McGuire P, Fusar-Poli P, Zandi MS, Lewis G, David AS. Psychiatric and neuropsychiatric presentations associated with severe coronavirus infections: a systematic review and meta-analysis with comparison to the COVID-19 pandemic. Lancet Psychiatry. 2020 Jul;7(7):611–627. doi: 10.1016/S2215-0366(20)30203-0. Epub 2020 May 18. PMID: 32437679; PMCID: PMC7234781.

16. Zhang P, Li J, Liu H, et al. Long-term bone and lung consequences associated with hospital-acquired severe acute respiratory syndrome: a 15-year follow-up from a prospective cohort study. Bone Res. 2020;8(1):8. [PMC free article] [PubMed] [Google Scholar]

17. Galanis P, Vraka I, Fragkou D, Bilali A, Kaitelidou D. Seroprevalence of SARS-CoV-2 antibodies and associated factors in healthcare workers: a systematic review and meta-analysis. J Hosp Infect. 2021;108:120–134. doi:10.1016/j.jhin.2020.11.008

18. Gómez-Ochoa SA, Franco OH, Rojas LZ, Raguindin PF, Roa-Díaz ZM, Wyssmann BM, Guevara SLR, Echeverría LE, Glisic M, Muka T. COVID-19 in Health-Care Workers: A Living Systematic Review and Meta-Analysis of Prevalence, Risk Factors, Clinical Characteristics, and Outcomes. Am J Epidemiol. 2021 Jan 4;190(1):161–175. doi: 10.1093/aje/kwaa191.

19. Müller SA, Wood RR, Hanefeld J, El-Bcheraoui C. Seroprevalence and Risk Factors of COVID-19 in Healthcare Workers From Eleven African Countries: A Scoping Review and Appraisal of Existing Evidence. Health Policy Plan. 2021 Nov 2:czab133. doi: 10.1093/heapol/czab133. Epub ahead of print. PMID: 34726740; PMCID: PMC8689910.

20. Sahu A.K., Amrithanand V.T., Mathew R., Aggarwal P., Nayer J., Bhoi S. COVID-19 in health care workers – a systematic review and meta-analysis. Am J Emerg Med. 2020;38:1727–1731.

21. Osikomaiya B, Erinoso O, Wright KO, et al. ’Long COVID’: persistent COVID-19 symptoms in survivors managed in Lagos State, Nigeria. BMC Infect Dis. 2021;21(1):304. Published 2021 Mar 25. doi:10.1186/s12879-020-05716-x

22. Groff D., Sun A., Ssentongo A.E., Ba D.M., Parsons N., Poudel G.R., Lekoubou A., Oh J.S., Ericson J.E., Ssentongo P., et al. Short-Term and Long-Term Rates of Postacute Sequelae of SARS-CoV-2 Infection: A Systematic Review. [(accessed on 8 December 2021)]; JAMA Netw. Open. 2021 4:e2128568. doi: 10.1001/jamanetworkopen.2021.28568.

23. Taquet M, Dercon Q, Luciano S, Geddes JR, Husain M, Harrison PJ. Incidence, co-occurrence, and evolution of long-COVID features: A 6-month retrospective cohort study of 273,618 survivors of COVID-19. PLoS Med. 2021;18(9):e1003773. Published 2021 Sep 28. doi:10.1371/journal.pmed.1003773

24. Hyassat D, El-Khateeb M, Dahbour A, Shunnaq S, Naji D, Bani Ata E. et al. Post-COVID-19 syndrome among healthcare workers in Jordan. East Mediterr Health J. 2023 Apr 27;29(4):247–253. doi: 10.26719/emhj.23.029.

25. Menges D, Ballouz T, Anagnostopoulos A, Aschmann HE, Domenghino A, Fehr JS, Puhan MA. Burden of post-COVID-19 syndrome and implications for healthcare service planning: A population-based cohort study. PLoS One. 2021 Jul 12;16(7):e0254523. doi: 10.1371/journal.pone.0254523. PMID: 34252157; PMCID: PMC8274847.

26. Davies, N., Klepac, P., Liu, Y., Prem, K., Jit, M., & Pearson, C. et al. (2020). Age-dependent effects in the transmission and control of COVID-19 epidemics. Nature Medicine, 26(8), 1205–1211. doi: 10.1038/s41591-020-0962-9.

27. Montenegro P, Moral I, Puy A, et al. Prevalence of Post COVID-19 Condition in Primary Care: A Cross Sectional Study. Int J Environ Res Public Health. 2022;19(3):1836. Published 2022 Feb 6. doi:10.3390/ijerph19031836

28. Iwu CJ, Iwu CD, Wiysonge CS. The occurrence of long COVID: a rapid review. Pan Afr Med J. 2021;38:65. Published 2021 Jan 20. doi:10.11604/pamj.2021.38.65.27366

29. Elnadi H, Odetokun IA, Bolarinwa O, Ahmed Z, Okechukwu O, Al-Mustapha AI (2020) Knowledge, attitude, and perceptions towards the 2019 Coronavirus Pandemic: A bi-national survey in Africa. PLoS ONE 15(7): e0236918. https://doi.org/10.1371/journal.pone.023691

30. Odetokun, I.A., Alhaji, N.B., Akpabio, U., Abdulkareem, M.A., Bilat, G.T., Subedi, D., Ghali-Mohammed, I. & Elelu, N. (2022): Knowledge, risk perception and prevention preparedness towards COVID-19 among a cross-section of animal health professionals in Nigeria. Pan African Medical Journal. 41(20).

31. Anjorin, A.A., Odetokun, I.A., Abioye, A.I., Elnadi, H., Umoren, M.V., Damaris, B.F., Eyedo, J., Umar, H.I., Nyandwi, J.B., Abdalla, M.M., Tijani, S.O., Awiagah, K.S., Idowu, G.A., Fabrice, S.A.N., Maisara, A.M.O., Razouqi, Y., Mhgoob, Z.E., Parker, S., Asowata, O.E., Adesanya, I.O., Obara, M.A., Jaumdally, S., Kitema, G.F., Okuneye, T.A., Mbanzulu, K.M., Daitoni, H., Hallie, E.F., Mosbah, R. & Fasina, F.O. (2021): Will Africans take COVID-19 vaccination? PLoS ONE. 16(12); e0260575.

32. Venkatesan, P. (2022). Do vaccines protect from long COVID?. The Lancet Respiratory Medicine, 10(3), e30. doi: 10.1016/s2213-2600(22)00020-0

33. Sheraton M, Deo N, Dutt T, Surani S, Hall-Flavin D, Kashyap R. Psychological effects of the COVID 19 pandemic on healthcare workers globally: A systematic review. Psychiatry research. 2020 Oct 1;292:113360.

34. Spoorthy MS, Pratapa SK, Mahant S. Mental health problems faced by healthcare workers due to the COVID-19 pandemic–A review. Asian journal of psychiatry. 2020 Jun 1;51:102119.

35. Tan BY, Chew NW, Lee GK, Jing M, Goh Y, Yeo LL, Zhang K, Chin HK, Ahmad A, Khan FA, Shanmugam GN. Psychological impact of the COVID-19 pandemic on health care workers in Singapore. Annals of internal medicine. 2020 Aug 18;173(4):317–20.

36. Temsah MH, Al-Sohime F, Alamro N, Al-Eyadhy A, Al-Hasan K, Jamal A, Al-Maglouth I, Aljamaan F, Al Amri M, Barry M, Al-Subaie S. The psychological impact of COVID-19 pandemic on health care workers in a MERS-CoV endemic country. Journal of infection and public health. 2020 Jun 1;13(6):877–82.

37. Vizheh M, Qorbani M, Arzaghi SM, Muhidin S, Javanmard Z, Esmaeili M. The mental health of healthcare workers in the COVID-19 pandemic: A systematic review. Journal of Diabetes & Metabolic Disorders. 2020 Dec;19(2):1967–78.

38. Yan L, Sun P, Wang M, Song T, Wu Y, Luo J, Chen L. The psychological impact of COVID-19 pandemic on health care workers: a systematic review and meta-analysis. Frontiers in psychology. 2021;12:2382.

39. Chersich MF, Gray G, Fairlie L, et al. COVID-19 in Africa: care and protection for frontline healthcare workers. Global Health. 2020;16(1):46. Published 2020 May 15. doi:10.1186/s12992-020-00574-3

40. Jalili M, Niroomand M, Hadavand F, Zeinali K, Fotouhi A. Burnout among healthcare professionals during COVID-19 pandemic: a cross-sectional study. Int Arch Occup Environ Health. 2021;94(6):1345–1352. doi:10.1007/s00420-021-01695-x

41. Torrente M, Sousa PA, Sánchez-Ramos A, et al. To burn-out or not to burn-out: a cross-sectional study in healthcare professionals in Spain during COVID-19 pandemic. BMJ Open. 2021;11(2):e044945. doi:10.1136/bmjopen-2020-044945

42. Salazar de Pablo G, Vaquerizo-Serrano J, Catalan A, et al. Impact of coronavirus syndromes on physical and mental health of health care workers: Systematic review and meta-analysis. J Affect Disord. 2020;275:48–57.

43. Schwartz RM, McCann-Pineo M, Bellehsen M, Singh V, Malhotra P, Rasul R, Corley SS, Jan S, Parashar N, George S, Yacht AC, Young JQ. The Impact of Physicians’ COVID-19 Pandemic Occupational Experiences on Mental Health. J Occup Environ Med. 2022 Feb 1;64(2):151–157. doi: 10.1097/JOM.0000000000002380

44. Chew NWS, Lee GKH, Tan BYQ, et al. A multinational, multicentre study on the psychological outcomes and associated physical symptoms amongst healthcare workers during COVID-19 outbreak. Brain Behav Immun. 2020;88:559–565. doi:10.1016/j.bbi.2020.04.049

45. Davis, H., Assaf, G., McCorkell, L., Wei, H., Low, R., & Re’em, Y. et al. (2021). Characterizing long COVID in an international cohort: 7 months of symptoms and their impact. Eclinicalmedicine, 38, 101019. doi: 10.1016/j.eclinm.2021.101019

46. Twycross A. Living with long Covid: some reflections 14 months down the line. Evid Based Nurs. 2021 May 26:ebnurs-2021-103449. doi: 10.1136/ebnurs-2021-103449

47. Loewenson R. COVID-19 in East and Southern Africa: Rebuilding Differently and Better Must Start Now. MEDICC Rev. 2020 Jul;22(3):59–60. doi: 10.37757/MR2020.V22.N3.13.

48. Mendelson M, Nel J, Blumberg L, Madhi SA, Dryden M, Stevens W, Venter FWD. Long-COVID: An evolving problem with an extensive impact. S Afr Med J. 2020 Nov 23;111(1):10–12. doi: 10.7196/SAMJ.2020.v111i11.15433.

49. Randremanana R, Lazoumar RH, Tejiokem MC, et al. Institut Pasteur International Network’s efforts to guide control measures against the coronavirus disease 2019 (COVID-19) epidemic among healthcare workers in Africa. Int J Infect Dis. 2021;103:525–526. doi:10.1016/j.ijid.2020.12.032

50. van Kessel SAM, Olde Hartman TC, Lucassen PLBJ, van Jaarsveld CHM. Post-acute and long-COVID-19 symptoms in patients with mild diseases: a systematic review. Fam Pract. 2022;39(1):159–167. doi:10.1093/fampra/cmab076

51. Hopman J, Allegranzi B, Mehtar S. Managing COVID-19 in Low- and Middle-Income Countries. JAMA. 2020;323(16):1549–1550. doi:10.1001/jama.2020.4169.

52. Lam MH. Mental morbidities and chronic fatigue in severe acute respiratory syndrome survivors: long-term follow-up. Arch Intern Med. 2009;169(22):2142–2147

53. Ngai JC, Ko FW, Ng SS, et al. The long-term impact of severe acute respiratory syndrome on pulmonary function, exercise capacity and health status. Respirology. 2010;15(3):543–550.

54. Aiash H, Khodor M, Shah J, et al. Integrated multidisciplinary post-COVID-19 care in Egypt [published correction appears in Lancet Glob Health. 2021 Jul;9(7):e915]. Lancet Glob Health. 2021;9(7):e908–e909. doi:10.1016/S2214-109X(21)00206-0

55. WHO. Keep health workers safe to keep patients safe: WHO [Internet]. Who.int. 2020 [cited 20 September 2022]. Available from: https://www.who.int/news/item/17-09-2020-keep-health-workers-safe-to-keep-patients-safe-who

